# Correlation Analysis Between Disease Severity and Inflammation-related Parameters in Patients with COVID-19 Pneumonia

**DOI:** 10.1101/2020.02.25.20025643

**Authors:** Jing Gong, Hui Dong, Qingsong Xia, Zhaoyi Huang, Dingkun Wang, Yan Zhao, Wenhua Liu, Shenghao Tu, Mingmin Zhang, Qi Wang, Fuer Lu

**Affiliations:** Department of Integrated Traditional Chinese and Western Medicine, Tongji Hospital, Tongji Medical College, Huazhong University of Science and Technology, Wuhan, China; Institute of Integrated Traditional Chinese and Western Medicine, Tongji Hospital, Tongji Medical College, Huazhong University of Science and Technology, Wuhan, China; Clinical Research Center, Tongji Hospital, Tongji Medical College, Huazhong University of Science and Technology, Wuhan, China

## Abstract

**Aim:** The new coronavirus (COVID-19) pneumonia outbreaking at the end of 2019 is highly contagious. Crude mortality rate reached 49% in critical patients. Inflammation matters on disease progression. This study analyzed blood inflammation indicators among mild, severe and critical patients, helping to identify severe or critical patients early.

**Methods:** In this cross-sectional study, 100 patients were included and divided to mild, severe or critical groups. Correlation of peripheral blood inflammation-related indicators with disease criticality was analyzed. Cut-off values for critically ill patients were speculated through the ROC curve.

**Results:** Significantly, disease severity was associated with age (R=-0.564, P<0.001), interleukin-2 receptor (IL2R) (R=-0.534, P<0.001), interleukin-6 (IL-6) (R=-0.535, P<0.001), interleukin-8 (IL-8) (R=-0.308, P<0.001), interleukin-10 (IL-10) (R=-0.422, P<0.001), tumor necrosis factor α (TNFα) (R=-0.322, P<0.001), C-reactive protein (CRP) (R=-0.604, P<0.001), ferroprotein (R=-0.508, P<0.001), procalcitonin (R=-0.650, P<0.001), white cell counts (WBC) (R=-0.54, P<0.001), lymphocyte counts (LC) (R=0.56, P<0.001), neutrophil count (NC) (R=-0.585, P<0.001) and eosinophil counts (EC) (R=0.299, P=0.01).

**Conclusion:** With following parameters such as age >67.5 years, IL2R >793.5U/mL, CRP >30.7ng/mL, ferroprotein >2252μg/L, WBC>9.5*10^9/L or NC >7.305*10^9/L, the progress of COVID-19 to critical stage should be closely observed and possibly prevented. Inflammation is closely related to severity of COVID-19, and IL-6, TNFα and IL-8 might be promising therapeutic targets.

## Introduction

Wuhan in China is gonging through a battle between humans and a virus named 2019 novel coronavirus (COVID-19). As far, it has infected more than 70000 people and over 2000 people were dead all across the country^[1]^. Phylogenetic analysis of viral complete genome revealed that the novel virus was most similar to a group of SARS-like coronaviruses (genus Betacoronavirus, subgenus Sarbecovirus) which stems from bats in China^[2]^. As one of the several coronavirus that are pathogenic to humans, including severe acute respiratory syndrome (SARS) coronavirus (SARS-CoV) and Middle East respiratory syndrome coronavirus (MERS-CoV)^[3]^, patients infected with COVID-19 have a series of clinical manifestations, including fever, cough, myalgia or fatigue, dyspnea, even acute respiratory distress syndrome(ARDS), acute cardiac injury and secondary infection, and a lot of severe patients had to been admitted to the intensive care unit (ICU)^[4]^.

Besides the positive viral nucleic acid analysis and the representative pulmonary CT findings (bilateral distribution of patchy shadows and ground glass opacity), most individual patients showed the changes of neutrophil count (NC), D-dimer, blood urea nitrogen, and creatinine levels and lymphocyte counts (LC)^[5]^. Increased inflammation-related biomarkers were found in patients with COVID-19 pneumonia, including C-reactive protein (CRP), ferroprotein, erythrocyte sedimentation rate (ESR) and interleukin-6 (IL-6)^[6]^. Furthermore, patients in ICU manifested higher cytokine levels of interleukin-2 (IL-2), interleukin-7 (IL-7), interleukin-10 (IL-10), granulocyte colony-stimulating factor (GSCF), interferon gamma-induced protein 10 (IP-10), chemokine (C-C motif) ligand 2 (CCL2), Chemokine (C-C motif) ligand 3 (CCL3), and tumor necrosis factor α (TNFα) than that of non-ICU patients^[4]^. However, the correlation between the inflammatory markers and the disease severity was not completely clear. Therefore, this study retrospectively analyzed blood inflammation indicators among mild, severe and critical patients, which may help to identify severe or critical patients early and perform clinical intervention early.

## Methods

### Patients

Test data of patients diagnosed with COVID-19 pneumonia whose peripheral blood cytokines were tested were collected in Tongji Hospital, Tongji Medical College of Huazhong university of Science and Technology for cross-sectional study. As listed below, the diagnosis of severe or critical patients were depending on “New Coronavirus-Infected Pneumonia” Severe and Critical Diagnosis and Treatment Program (Second trial version) formulated by the National Health Commission of China.

### Standard of severe or critical patients

Severe patients should have any of the following conditions:

1. Respiratory distress, RR ≥30 times / minute;
2. Under the resting state, the oxygen saturation ≤93%;
3. Oxygen partial pressure (PaO2)/oxygen concentration (FiO2) in arterial blood ≤300mmHg.
4. >50% lung imaging progress in the short term.

Critical patients should have any of the following conditions:

1. Respiratory failure occurs and mechanical ventilation required;
2. Shock occurs;
3. Combining other organ failure and requiring treatment in ICU.

### Inflammation-related biomarkers

All tests were completed in the clinical laboratory in Tongji Hospital. Interleukin-1β (IL-1β), interleukin-2 receptor (IL2R), interleukin-8 (IL-8), IL-10 and TNFα were detected by Siemens chemiluminescence method and IL-6 were detected by Roche electrochemiluminescence method according to the manufacturer’s instruction. The ultrasensitive CRP regent was provided by Nippon Denkasei Co., Ltd, and CRP was detected by immunoturbidimetry method. Ferroprotein was detected by Roche granule-enhanced immune turbidimetry. Procalcitonin (PCT) was detected by Roche electrochemiluminescence method. ESR was measured by Westergren’s international standard method. Peripheral blood cell was detected by fluorescence staining flow cytometry, and we analyzed the differences of white blood cell (WBC), NC, LC and eosinophils (EC) among three groups.

### Statistical analysis

For data of normal distribution (IL2R, ESR, ferroprotein, WBC and NC), comparisons among critical, severe and mild groups were analyzed by ANOVA analysis. With different homogeneity of variance, the pairwise comparison between groups was performed using the Bonferroni test or Dunnett’s T3 test. For non-normal distribution (CRP and LC), the data were conversed using square root, followed by ANOVA analysis and pairwise comparison. With data below the detectable limit, including IL-1β, IL-6, IL-8, IL-10, TNFα, PCT, and EC, the data were ranked referring to the reference range and value rank (Table 1); then non-parametric Kruskal-Wallis test were performed. In correlation analysis, Spearman correlation coefficient was used for the variables of normal distribution, Pearson correlation coefficient for those of skewed distribution, and Kendall’s tau-b correlation coefficient for ranked data.

**Table 1.** Reference range and analytical levels of inflammation-related test items.

| Test item | Reference range | analytical levels |  |  |  |
| --- | --- | --- | --- | --- | --- |
|  |  | 1 | 2 | 3 | 4 |
| **IL-2R** | 223-710 (U/mL) |  | - |  |  |
| **IL-6** | <7 (pg/mL) | <7 | 7-30 | 31-100 | >100 |
| **IL-8** | <62 (pg/mL) | 0-31 | 31-62 | >62 | - |
| **IL-10** | <9.1 (pg/mL) | < 5 | 5-9.1 | 9.1-20 | >20 |
| **TNF-α** | <8.1 (pg/mL) | <8.1 | 8.1-11 | >11 | - |
| **IL-1β** | <5 (pg/mL) | <5 | $\geq 5$ | 1 | |
| **CRP** | <1mg/L |  | 1 |  |  |
| **Ferroprotein** | Male: 30-400 (μg/mL)<br>Female: 15-150(μg/mL) | | - | | |
| **Procalcitonin** | 0.02-0.05ng/mL | <0.05 | 0.05-0.1 | 0.1-1 | >1 |
| **ESR** | 0-20mm/H |  | - |  |  |
| **WBC** | $3.5-9.5 \times 10^9/L$ | | - | | |
| **NC** | $1.8-6.3 \times 10^9/L$ | | - | | |
| **LC** | $1.1-3.2 \times 10^9/L$ | | - | | |
| **EC** | $0.02-0.05 \times 10^9/L$ | >0.1 | 0.02-0.1 | 0-0.02 | 0 |

To find out cut-off points of inflammatory parameters for critical patients, receiver operating characteristic (ROC) analyses were administrated. AUC was used for prediction strength and optimum cut-off points were chosen using Youden’s index. Data were analyzed using SPSS 20.0.

#### Ethics approval

This study was approved by the ethical committee of Tongji Hospital, Tongji Medical College, Huazhong University of Science and Technology. Because of the infectivity and the exploration urgency for this disease, written informed consent was waived by the ethical commission of Tongji Hospital.

## Results

Among the 100 included patients, 34 patients belong to mild group, 34 were severe, and 32 were critical. The average age was 57.02 years, and 59% patients were male. As shown in Figure 1A, the age of mild group (mean±SD: 45.29±13.08 years) was significantly different from that of severe patients (mean±SD: 60.41±9.80 years) or critical patients (mean±SD: 65.88±13.61 years), and there was no significant difference between severe and critical patients. To better detect the critical illness, the ROC curve of age was administrated and listed in Figure 2A (AUC=0.755, P =0.000). The best cut-off point of age was 67.5 years with a specificity of 88.2% and a sensitivity of 59.4%. The mean IL2R level was 1317.31 U/mL in critical group, 885.09 U/mL in severe group and 486.44 U/mL in mild group, with significant differences among three groups (Figure 1B). The ROC curve of IL2R (AUC=0.8, P=0.000) was shown in Figure 2B, and the best cut-off point was 793.5 U/mL with a specificity of 67.6% and a sensitivity of 84.4%.

**Figure 1.**
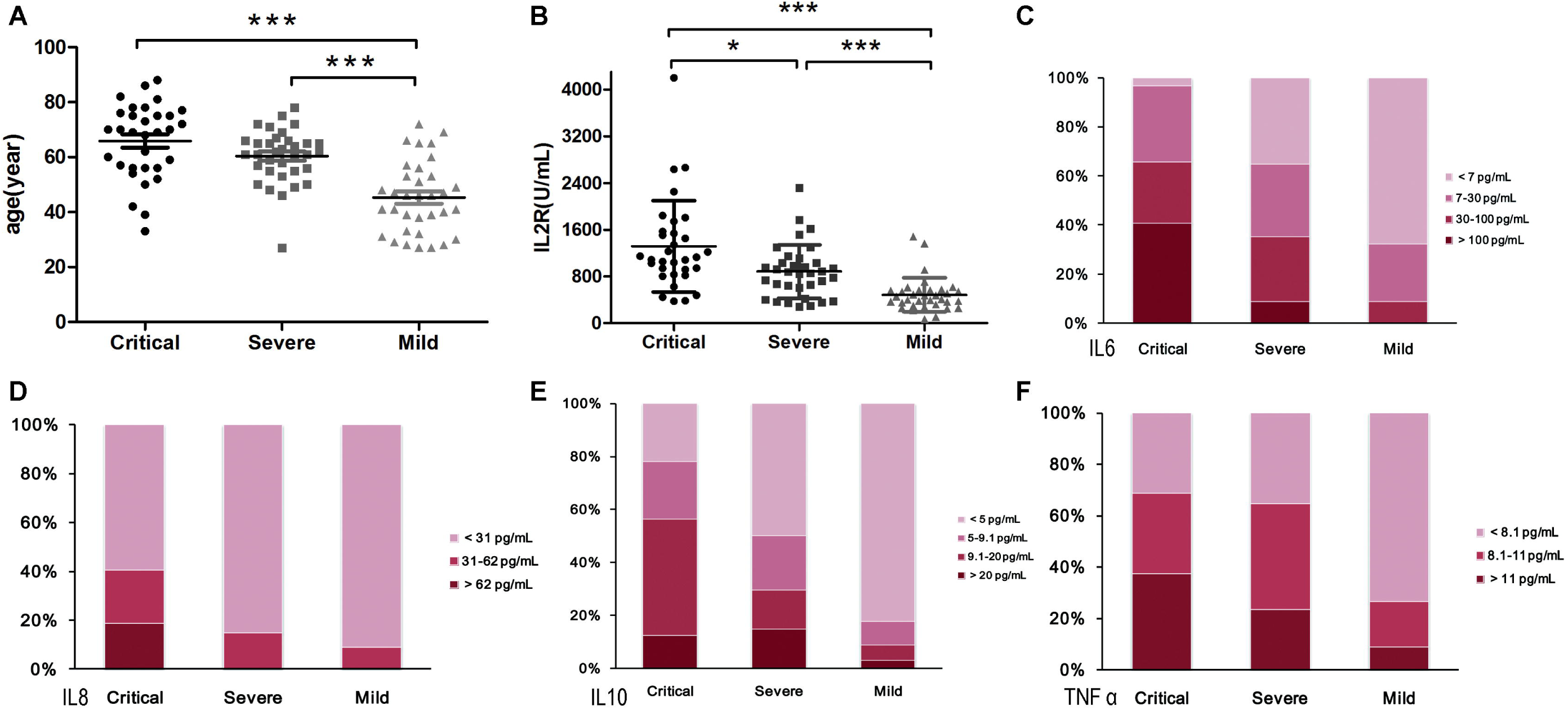
Comparison of age and cytokines among mild, severe and critical patients with COVID-19 pneumonia. Data are presented as mean ± standard error or percentage. (A) age; (B)IL2R; (C) IL-6; (D) IL-8; (E) IL-10; (F) TNFα. *P<0.05; ***P<0.001.

**Figure 2.**
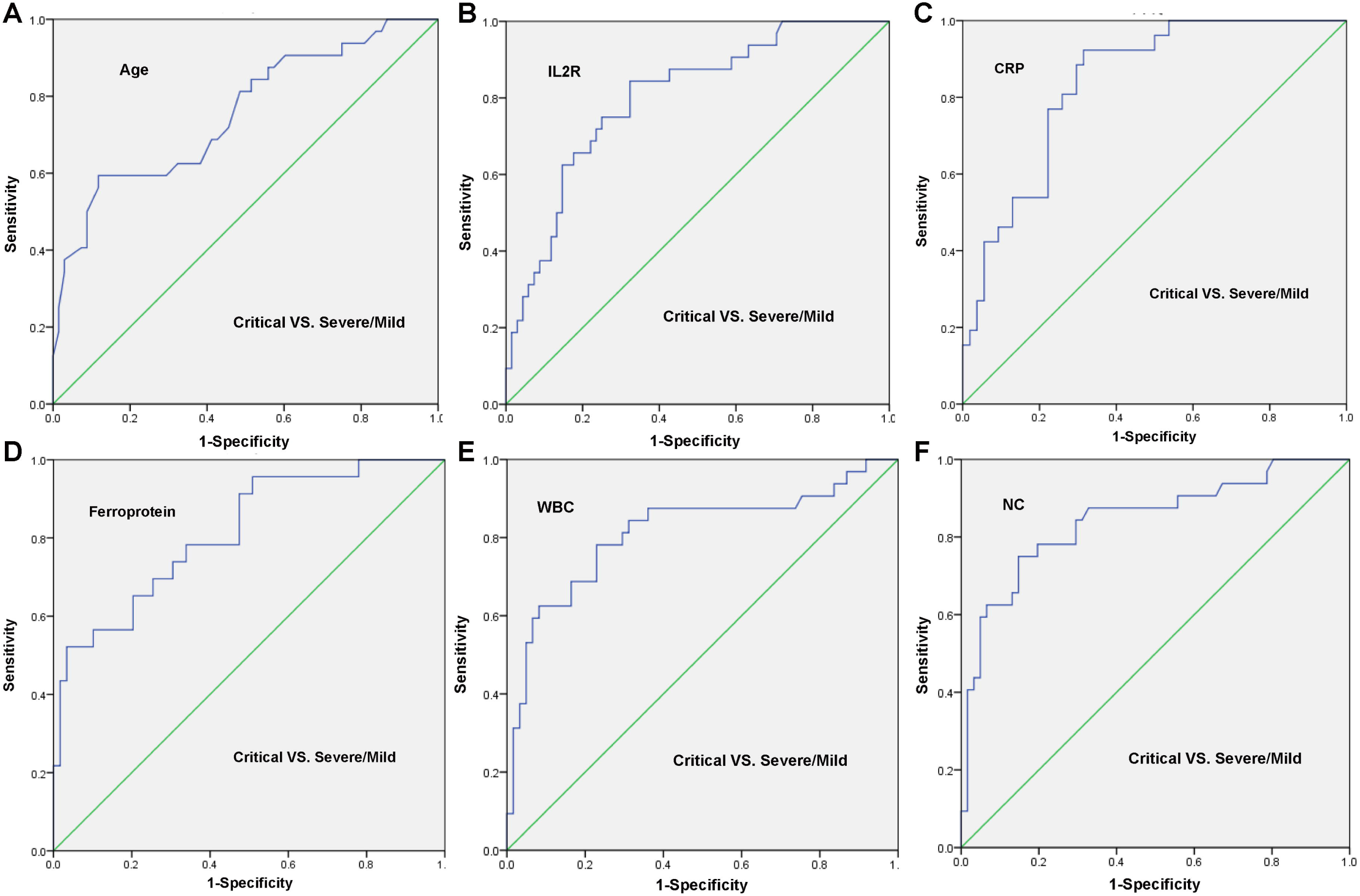
ROC curve of age and inflammatory parameters. (A) age; (B)IL2R; (C) CRP; (D) ferroprotein; (E) WBC; (F) NC.

Because some data were below the detectable limit, the test values were graded according to reference range and rank order regarding IL-6, IL-8, IL-10 and TNFα (Table 1). As shown in Figure 1C and Table 2, the IL-6 levels were significantly different among three groups, and all IL-6 concentration was <100 pg/mL in mild patients. Although the IL-8 reference range was <62 pg/mL, the IL-8 levels of most patients were <31 pg/mL, three ranked grades were set, namely, <31 pg/mL, 31-62 pg/mL and >62 pg/mL. Figure 1D and Table 2 showed that all IL-8 levels in mild and severe patients were within the reference range (<62 pg/mL), and there were significant differences between critical and severe patients or critical and mild group. With regard to IL-10 and TNFα, there was significant difference between mild and severe or mild and critical patients, and no significant difference was found between severe and critical group (Figure 1E-F). With regard to cytokine IL-1β, no significant difference was found among three groups, and the data were not shown.

**Table 2.** Statistical differences in analysis of ranked data.

|  | Critical-Severe | Critical-Mild | Severe-Mild |
| --- | --- | --- | --- |
| **IL-6** | ** | *** | * |
| **IL-8** | * | ** | - |
| **IL-10** | - | *** | * |
| **TNFα** | - | ** | * |
| **IL-1β** | - | - | - |
| **Procalcitonin** | ** | *** | ** |
| **EC** | - | ** | * |

In addition, levels of CRP, ferroprotein and PCT were statistically different among mild, severe and critical patients (Figure 3A-C). All PCT levels were <0.1 ng/mL in the mild patients, while PCT concentrations in all critical patients were >0.05 ng/mL. ROC curve of CRP (AUC=0.838, p =0.000, Figure 2C) suggested the best cut-off point was 30.7 ng/mL with a specificity of 92.3% and a sensitivity of 68.5%. The average values of ferroprotein were 2753.87μg/L in critical group, 1147.55μg/L in severe group and 475.85μg/L in mild group, respectively. ROC curve of ferroprotein (AUC=0.814, P=0.000, Figure 2D) indicated the best cut-off point was 2252μg/L with a specificity of 96.6% and a sensitivity of 52.2%. There was no statistical difference on ESR among three groups (Figure 3D).

**Figure 3.**
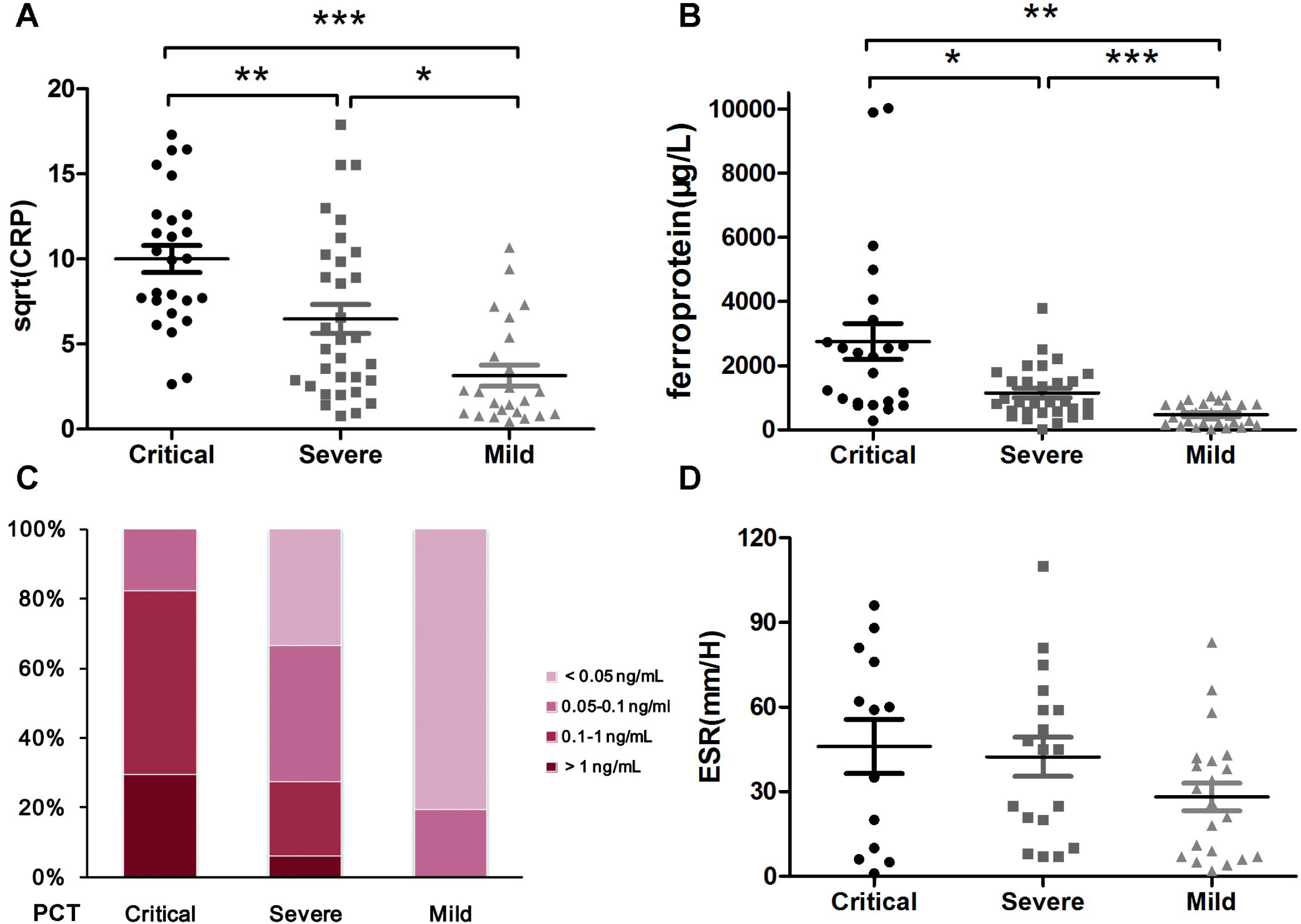
Comparison of inflammatory parametes among mild, severe and critical patients with COVID-19 pneumonia. (A) sqrt(CRP); (B) ferroprotein; (C) PCT; (D) ESR. Data are presented as mean ± standard error or percentage. *P<0.05; **P<0.05; ***P<0.001.

In peripheral blood cell analysis, there were significant differences in WBC count between the critical and severe groups or critical and mild groups, and no significance was found between the mild and severe groups (Figure 4A). ROC curve of WBC (AUC =0.838, p =0.000, Figure 2E) suggested the best cut-off point was 7.92*10^9/L with 77% specificity and 78.1% sensitivity. If the detected values of WBC are above the upper reference limit, it often indicates infection. Because WBC count is influenced by therapy, clinically the upper limit of reference range of WBC, namely 9.5*10^9/L, seems more meaningful.

**Figure 4.**
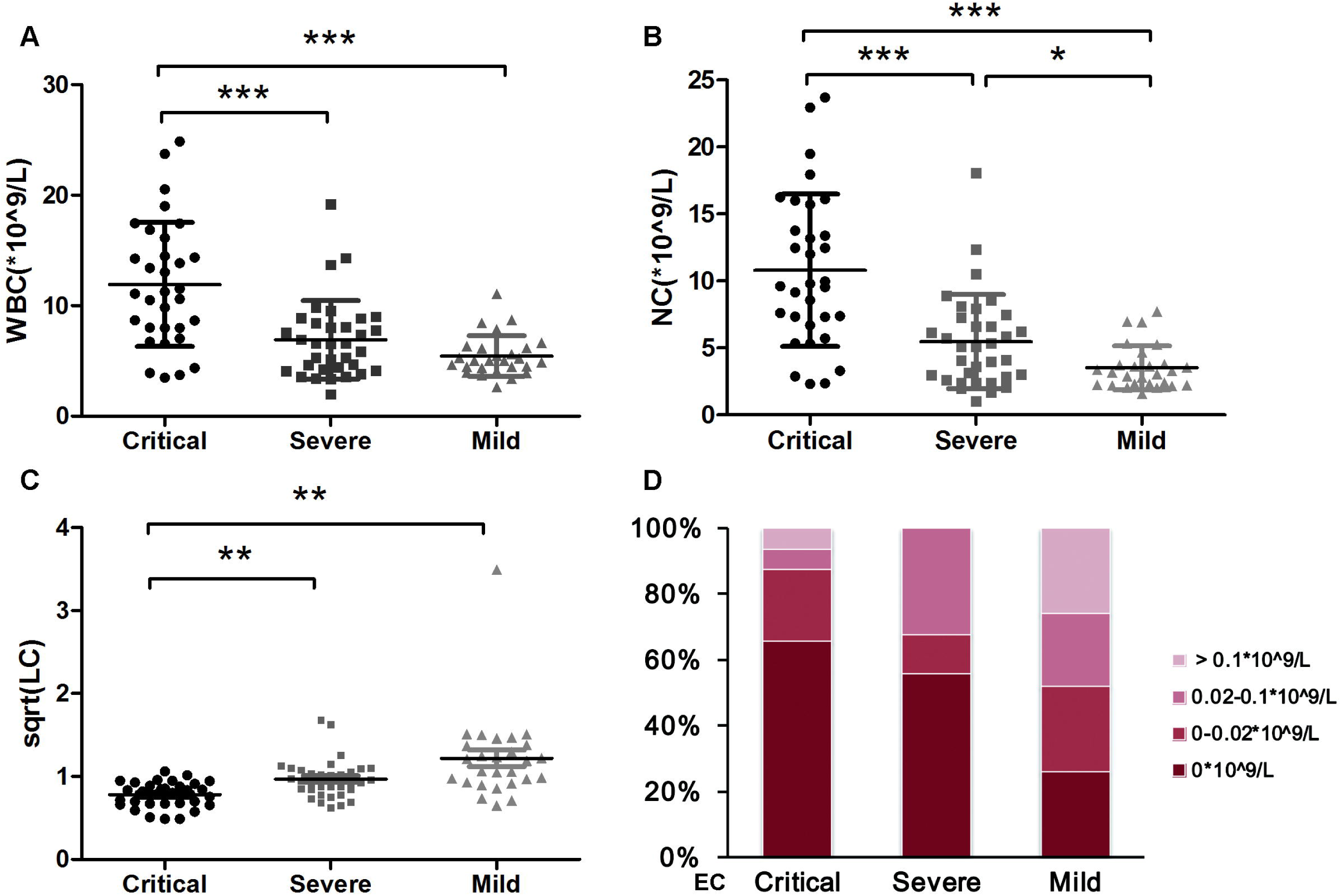
Comparison of blood cell counts among mild, severe and critical patients with COVID-19 pneumonia. Data are presented as mean ± standard error or percentage. (A) WBC; (B) NC; (C) sqrt(LC); (D) EC.

NC was significantly different among the three groups (Figure 4B), and the average was 10.80*10^9/L in critical group, 5.47*10^9/L in severe group and 3.53 *10^9/L in mild group. ROC curve of NC (AUC=0.814, P=0.000, Figure 2F) indicated the best cut-off point was 7.305*10^9/L with 85.2% specificity and 75% sensitivity. There was significantly difference about LC between critical and severe group or critical and mild group (Figure 4C). Many patients in each group had a decreased level of EC, and 0 EC was detected in 65.63% patients in critical group, 55.88% in severe group, and 25.93% in mild group. There were significant differences between severe and mild or critical and mild patients (Table 2, Figure 4D).

Table 3 showed correlation analysis results between the indicators with disease severity. Significant correlations were found about age, IL2R, IL-6, IL-8, IL-10, TNFα, CRP, ferroprotein, PCT, LC, NC, and EC. PCT (R=-0.650), CRP (R=-0.604), NC (R=-0.585), age (R=-0.564), LC (R=0.56), IL-6 (R=-0.535) IL2R(R=-0.534), and ferroprotein (R=-0.508) were highly correlated (Table 3). There was no significant correlation on IL-1β, gender and ESR.

**Table 3.** Correlation coefficient and P value between items and disease severity.

|  | **R** | **P** |
| --- | --- | --- |
| **age** | -0.564 | 0.000 |
| **gender** | 1.0 | 0.291 |
| **IL-2R** | -0.534 | 0.000 |
| **IL-6** | -0.535 | 0.000 |
| **IL-8** | -0.308 | 0.01 |
| **IL-10** | -0.422 | 0.000 |
| **TNF-α** | -0.322 | 0.000 |
| **IL-1β** | 0.098 | 0.301 |
| **CRP** | -0.604 | 0.000 |
| **Ferroprotein** | -0.508 | 0.000 |
| **Procalcitonin** | -0.650 | 0.000 |
| **ESR** | -0.261 | 0.061 |
| **WBC** | -0.54 | 0.000 |
| **NC** | -0.585 | 0.000 |
| **LC** | 0.56 | 0.000 |
| **EC** | 0.299 | 0.01 |

## Discussion

The new coronavirus (COVID-19) pneumonia outbreaking at the end of 2019 is highly contagious, with a crude mortality rate of about 2.3%^[1]^. Currently, more than 74677 patients have been reported in the Mainland of China. About 80.9% patients are with mild to moderate severity^[1]^, and with a better prognosis. However, for patients developing into severe or critical levels, the mortality rate was significantly increased, and crude mortality rate reached 49% in critical patients^[1]^. It plays key roles to identify severe and critical patients even earlier, aiming to improve the recovery rate and reduce mortality.

Clinically, many patients appeared short-term progressive aggravation, scholars speculated that “inflammatory storms” occurred, namely, overreaction of cytokine. In a preprint article in medrxiv, the lymphocyte subsets and cytokines of 123 patients (102 mild and 21 severe patients) were analyzed and the researchers found the numbers of CD4+ T cells and CD8+ T cells decreased, and the levels of IL-6 and IL-10 increased in severe cases^[7]^. In this study, a retrospective analysis was conducted about cytokines and other inflammatory indicators of 100 patients (including 66 severe or critical patients), and more indicators were expected to better identify critical patients and help clinical decision-making.

In accordance with Wan S’s^[7]^ and Liu J’s^[8]^ study, this study also found that the levels of IL-6 and IL-10 were associated with the severity of COVID-19 pneumonia; similar to Chaolin Huang’s study^[4]^, TNFα concentration, NC count and LC count were also found correlated with disease severity. In addition, IL2R levels, ferroprotein levels, PCT levels, and EC counts were also related to the disease severity. Besides, we also found the IL-6 levels in mild patients were lower than 100pg/mL. For eight patients who were deceased in critical group, IL-6 level was >100pg/mL in three persons. IL-6 >100pg/mL might represent the emergence of “inflammatory storm”. IL-8 levels in mild and severe patients were normal, and for patients with IL-8 >62 pg/mL, more attention need to avoid the disease progression. Based on clinical practice and ROC analysis between critical and non-critical patients, some cut-off values of the test items were obtained. With age >67.5 years, IL2R >793.5U/mL, CRP >30.7ng/mL, ferroprotein >2252μg/L, WBC >9.5*10^9/L or NC>7.305*10^9/L, progress to critical illness should be closely observed and prevented.

Preventing recognition or blocking the occurrence of inflammatory action, new drugs development on immune regulation, might be new breakthroughs in the control of COVID-19 pneumonia. Pathological examination found lymphocyte-dominated mononuclear cell infiltration in interstitial pulmonary tissue, CD4+ and CD8+ T cells were significantly reduced in peripheral blood cells, but a high ratio of CD38+ (CD4 3.47%) and HLA-DR+ (CD8 39.4%) T cells^[9]^, which suggested excessive activation of pro-inflammatory cells. Presumably, lymphocyte deposition in lung tissue might contribute to lymphocyte reduction in peripheral blood. In addition, researchers found that CCR4+ CCR6+ Th17 cells were increased, and the IL-17 inhibitor (Secukinumab) against activated Th17 cells is promising for disease control^[9]^. In this study, IL-6, TNFα and IL-8 may be potential targets for immunotherapy of COVID-19. With IL-6 >100pg/mL, the patient’s condition was extremely critical. According to the news, a research team from First Affiliated Hospital of University of Science and Technology of China has used IL-6 receptor recombinant monoclonal antibody, Tozhu monoclonal antibody, in 14 critically or severely ill COVID-19 patients^[10]^. The results were reported to be encouraging. Respiratory oxygenation indices of 14 patients have improved to varying degrees, suggesting the potential of blocking inflammatory progression to reduce mortality^[10]^. IL-10 and IL2R levels were also related to disease severity, but they mainly inhibit the inflammatory response. Does it mean the simultaneous inflammatory and the anti-inflammatory reaction? The role of immunosuppression in disease progression and whether IL-10 and IL2R are possible therapeutic targets remains to be studied.

No significant difference was found about reduction ratio of B lymphocyte in the Wan S’s study^[7]^. In this study, there was no significant difference about Ig A, Ig G, IgM, C3 and C4 concentrations, suggesting humoral immunity did not appear to play a key role in disease progression. The role of humoral immunity in the recovery of COVID-19 needs further study.

Severe bacterial, fungal and parasitic infections, sepsis, systemic inflammatory response syndrome, as well as multiple organ dysfunction syndrome, PCT levels in serum are elevated, while PCT is generally not elevated with virus infections^[11]^. In our study, PCT concentrations in all critical patients were >0.05 ng/mL, and it suggested the possibility of multiple infections in critically-ill patients and the necessity of rational use of antibiotics. With elevated WBC and PCT, multiple infections may occur; without increased PCT, the increase of WBC and NC might be induced by glucocorticoids.

In clinical practice, few young and middle-aged women without basic disease progressed to critically illness, but our study did not find a gender correlation. Besides, Different from Chen N’s study^[6]^, no significant difference in ESR was found among three groups. One defect of the study is the limited sample size, and the conclusion needs to be further supported. Besides, glucocorticoids have effects on WBC and NC counts and the treatment procedure was not included in the analysis, the statistical differences of WBC and NC need further verification. In the next study, more large sample size, combined analysis with basic diseases, coagulation function, myocardial enzymes, liver and kidney function, and the type, dosage and time of medication need to identify the critical disease progression factors and better medication.

## Data Availability

The datasets generated and analyzed during the current study are available from the corresponding author on reasonable request.

## Acknowledgement

Thanks to all people who work so hard to fight against the COVID-19 pneumonia.

## References

[1] The Novel Coronavirus Pneumonia Emergency Response Epidemiology Tanm, The epidemiological characteristics of an outbreak of 2019 novel coronavirus diseases (covid-19) in china [J]. Zhonghua Liu Xing Bing Xue Za Zhi, 2020, 41(2):145–151.

[2] Wu F, Zhao S, Yu B, et al. A new coronavirus associated with human respiratory disease in china [J]. Nature, 2020, 3(10):020–2008.

[3] Lu R, Zhao X, Li J, et al. Genomic characterisation and epidemiology of 2019 novel coronavirus: Implications for virus origins and receptor binding [J]. Lancet, 2020, 30(20):30251–30258.

[4] Huang C, Wang Y, Li X, et al. Clinical features of patients infected with 2019 novel coronavirus in wuhan, china [J]. Lancet, 2020, 395(10223):497–506.

[5] Wang D, Hu B, Hu C, et al. Clinical characteristics of 138 hospitalized patients with 2019 novel coronavirus-infected pneumonia in wuhan, china [J]. Jama, 2020,7(2761044).

[6] Chen N, Zhou M, Dong X, et al. Epidemiological and clinical characteristics of 99 cases of 2019 novel coronavirus pneumonia in wuhan, china: A descriptive study [J]. Lancet, 2020, 395(10223):507–513.

[7] Wan S, Yi Q, Fan S, et al. Characteristics of lymphocyte subsets and cytokines in peripheral blood of 123 hospitalized 2 patients with 2019 novel coronavirus pneumonia (NCP), medRxiv preprint 2020, doi: https://doi.org/10.1101/2020.02.16.20023671.

[8] Liu J, Li S, Liu J, et al. Longitudinal characteristics of lymphocyte responses and cytokine profiles in the peripheral blood of SARS-CoV-2 infected patients, medRxiv preprint 2020, doi: https://doi.org/10.1101/2020.02.16.20023671.

[9] Xu Z, Shi L, Wang Y, et al. Pathological findings of COVID-19 associated with acute respiratory distress syndrome[J]. Lancet 2020, Doi : https://doi.org/10.1016/S2213-2600(20)30076-X

[10] https://mp.weixin.qq.com/s/b8v40pNb1H3TkzlOeLoysw

[11] Albrich WC, Harbarth S. Pros and cons of using biomarkers versus clinical decisions in start and stop decisions for antibiotics in the critical care setting [J]. Intensive Care Med, 2015, 41(10):1739–1751.

